# Vaccine efficacy trials for Crimean-Congo haemorrhagic fever: insights from modelling different epidemiological settings

**DOI:** 10.1101/2022.06.09.22276201

**Authors:** Juan F Vesga, Raphaёlle Métras, Madeleine H A Clark, Edris Ayazi, Andrea Apolloni, Toby Leslie, Veerle Msimang, Peter N. Thompson, W John Edmunds

**Affiliations:** Centre for Mathematical Modelling of Infectious Diseases, London School of Hygiene & Tropical Medicine, London, UK; Department of Infectious Disease Epidemiology, London School of Hygiene & Tropical Medicine, London, UK; INSERM, Sorbonne Université, Institut Pierre Louis d’Épidémiologie et de Santé Publique (Unité Mixte de Recherche en Santé 1136), Paris, France; Integrated Understanding of Health, Research Strategy and Programmes, Biotechnology and Biosciences Research Council, Swindon, UK; Ministry of Public Health, Massoud Square, Kabul, Afghanistan; CIRAD, UMR ASTRE, Montpellier, France; ASTRE, Univ Montpellier, CIRAD, INRA, Montpellier, France; International Health, London, UK; Epidemiology Section, Department of Production Animal Studies, Faculty of Veterinary Science, University of Pretoria, Onderstepoort, South Africa; Centre for Emerging Zoonotic and Parasitic Diseases, National Institute for Communicable Diseases of the National Health Laboratory Service, Sandringham, South Africa

**Author notes:** corresponding author: J F Vesga. **Funding** W.J.E., JFV and MHAC were funded by the Department of Health and Social Care using UK Aid funding managed by the National Institute for Health Research (Vaccine Efficacy Evaluation for Priority Emerging Diseases: PR-OD-1017-20002). The views expressed in this publication are those of the author(s) and not necessarily those of the Department of Health and Social Care.

**Keywords:** Crimean-Congo haemorrhagic fever, vaccines, mathematical modelling, clinical trials

## Abstract

**Background:** Crimean-Congo haemorrhagic fever (CCHF) is a priority emerging pathogen for which a licensed vaccine is not yet available. We aim to assess the feasibility of conducting phase III vaccine efficacy trials and the role of varying transmission dynamics.

**Methods:** We calibrate models of CCHF virus (CCHFV) transmission among livestock and spillover to humans in endemic areas in Afghanistan, Turkey and South Africa. We propose an individual randomised controlled trial targeted to high-risk population, and use the calibrated models to simulate trial cohorts to estimate the minimum trial endpoints necessary to analyse vaccine efficacy, sample size and follow-up time in the three settings.

**Results:** Under assumptions of a minimum vaccine efficacy of 60%, the minimum sample size needed to accrue the required 150 clinical endpoints in a minimum follow-up time of 6 months is estimated to be 34,000 (CrI 95%, 16,750 – 88,725) and 37,000 (CrI 95%, 13,000 – 77,250) in Afghanistan and Turkey, respectively. The results suggest that for South Africa the low endemic transmission levels will not permit achieving the necessary conditions for conducting this trial within a realistic follow-up time. In a scenario of CCHFV infection (rather than clinical case) as trial endpoint, the required sample size is reduced by 70% to 80% in Afghanistan and Turkey, and in South Africa, a trial becomes feasible for large sample sizes (>75,000) and vaccine efficacy of >70%. Increased expected vaccine efficacy >60% will reduce the required number of trial endpoints and thus the sample size and follow-time in phase III trials.

**Conclusions:** Underlying endemic transmission levels will play a central role in defining the feasibility of phase III vaccine efficacy trials. Endemic settings in Afghanistan and Turkey offer conditions under which such studies could feasibly be conducted.

## Introduction

Crimean-Congo haemorrhagic fever (CCHF) virus (CCHFV) is a zoonotic tick-borne emerging pathogen that can lead to fatal haemorrhagic fever in humans. Several CCHFV vaccine candidates are under different phases of study, including inactivated virus [1,2], DNA [3], mRNA [4], and plant-expressed glycoprotein formulations [5], amongst others. Despite this, no effective vaccine formulation is currently available, while the epidemiological map of influence for CCHFV keeps expanding [6–8]. Recognising this urgency, WHO has included CCHFV as one of the emerging pathogens which requires accelerated efforts to develop improved diagnostics, therapeutics and effective and safe vaccines [9]. This last point is particularly challenging as several factors can hinder the design and performance of vaccine efficacy trials for emerging infections. The most evident one is the limited commercial incentive for investing in costly randomised trials for “low burden” and context-specific zoonoses, but equally important are the difficulties intrinsic to trial design when the expected volume of clinical cases is highly uncertain or low.

Findings from our previous analysis of the transmission dynamics of CCHFV in an endemic region in Afghanistan [10] suggest that a future vaccination campaign targeted to human groups at high risk of infection should be preferred over animal vaccination in this setting. Here we examine the feasibility of conducting a phase III vaccine trial for a CCHFV-specific vaccine, by estimating key trial design components like sample size and time to attain the necessary number of trial endpoints, in three different endemic settings: Afghanistan, South Africa and Turkey.

## Methods

### Study locations and data

For this study, we select endemic areas in three different countries, namely, Herat in Afghanistan, Free State, North West and Northern Cape provinces in South Africa and, Tokat, Sivas, Erzurum, Erzincan and Gümüşhane provinces in the Kelkit valley in Turkey (**Fig. 1**).

**Figure 1.**
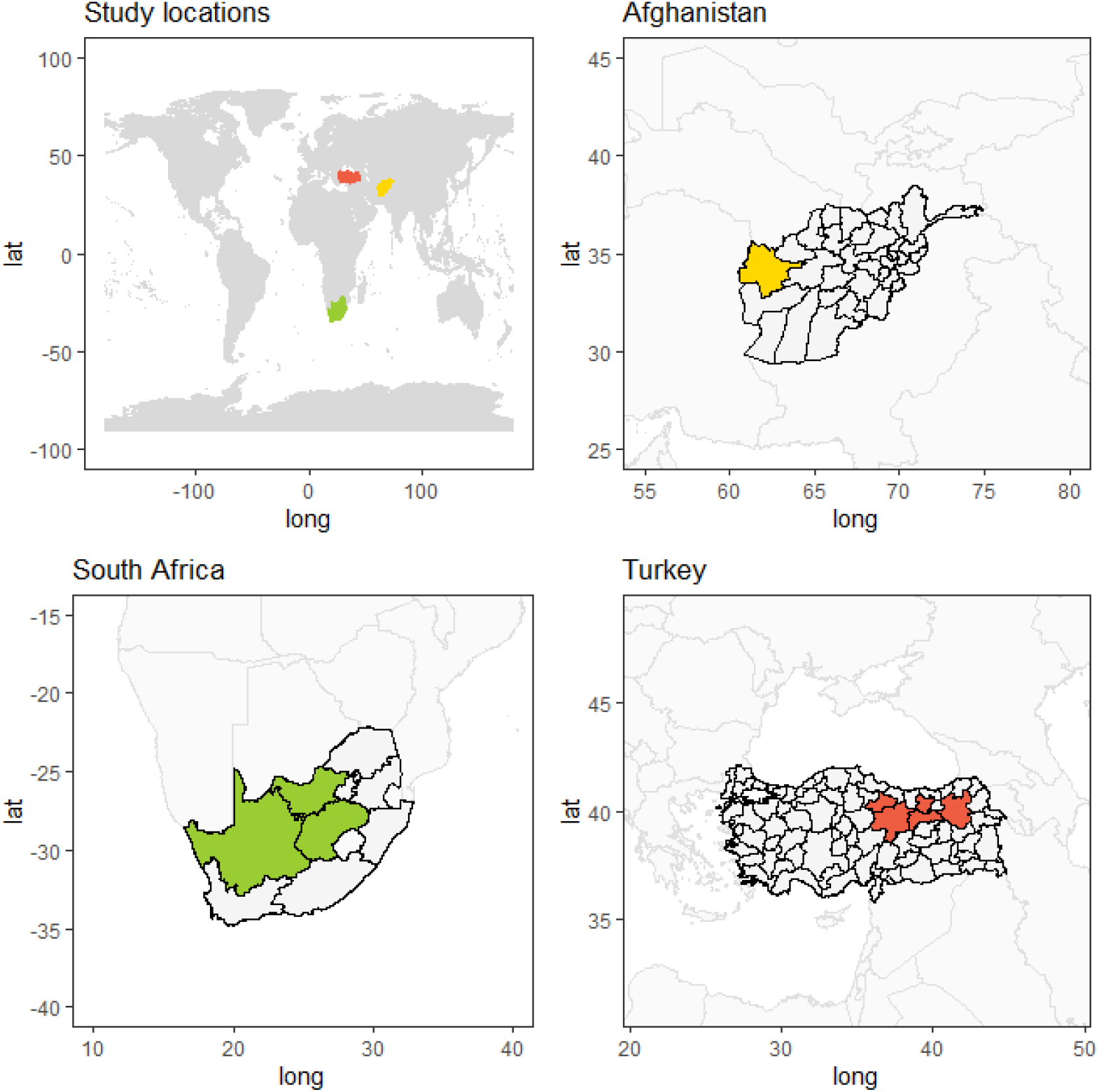
Geographical locations in the study. Data from endemic provinces on CCHF incidence and seroprevalence in humans and animals. In Afghanistan, we simulate and use epidemiological and environmental data from Herat (yellow). In South Africa we considered the Free State, North West and Northern Cape provinces (green), while in Turkey we model Tokat, Sivas, Erzurum, Erzincan and Gümüşhane provinces in the Kelkit valley.

In Afghanistan, the first case of CCHF was reported in 1998, and then intermittently until the start of national active surveillance for CCHFV in 2007 [11]. In this context at least two major CCHF outbreaks have been characterised in Herat, western Afghanistan, one in 2008 [12], and more recently in 2017 [13], with reported case fatality rates of 33% and 22% respectively. In this same area of the country, high levels of IgG antibodies have been identified in livestock, suggesting sustained endemic transmission in animal hosts [14]. Here we use animal and human seroprevalence data, and reports of clinical human cases from Herat, as described in the supporting information **Table S1**.

The first cases of CCHF were reported as early as 1981 in South Africa in the North West Province, and since then sporadic cases have occurred annually, with 217 cases reported between 1981-2020, with most cases coming from the North West, Northern Cape, Free State and Western Cape provinces [15]. Different cross-sectional studies over the years have shown a high seroprevalence in livestock (32%-80%) [16], and more recently a study from the Free State and Northern Cape provinces reported a prevalence of 74.2% (95%CI: 64.2–82.1%) in cattle and 3.9% (95%CI: 2.6–5.8%) in farm and wildlife workers [17]. For South Africa we calibrated our models to data reflecting human and animal seroprevalence and human cases from three provinces in South Africa: Free State, North West and Northern Cape provinces.

In Turkey, the first cases of CCHF were identified in 2002 in Tokat city, Anatolia, amongst people involved in farming and animal husbandry [18]. A number of seroprevalence studies in humans and animals across the country have helped to identify Kelkit valley in the northern part of the country as an endemic hotspot for CCHFV[19]. Case reports reached a peak between 2008 and 2009 when ∼1,300 cases were confirmed each year. Since then, around 900 cases are reported yearly [18]. Importantly, since 2003 Turkey has established a country-wide surveillance system and has improved its reporting capacity for CCHF, specifically increasing the number of reference laboratories with the capacity to perform ELISA for IgM and IgG and RT-PCR. This is thought to explain why Turkey has so many more reported cases annually than any country in the region [20]. In 2021, 243 cases had been reported by July in Turkey and 13 fatalities attributed to CCHF [21].

For Turkey we capture these trends using seroprevalence data in animals and humans and human clinical cases reported from the five provinces reporting over 70% of CCHFV cases every year, namely Tokat, Sivas, Erzurum, Erzincan, and Gümüşhane.

Our country selection reflects a spectrum in incidence of CCHF cases reported, in which, as described above, South Africa lies at the lowest bound, Turkey in the upper bound and Afghanistan in the middle. This provides us with a wide epidemiological context for testing vaccine trial feasibility.

Data from the three countries are summarised in the supporting information **Table S1**.

### CCHFV transmission model

We model transmission of CCHFV in livestock and spillover into humans with a combined modelling framework. Briefly, for livestock we use an age-structured deterministic Susceptible-Infectious-Recovered-Susceptible (SIRS) design in which we introduce selected environmental drivers as means to capture the environment-dependant seasonality in tick activity, in the absence of tick data in these locations. We test four potential drivers of tick activity, namely, soil temperature, saturation deficit, normalized difference vegetation index (NDVI), and relative humidity. For each country we use DIC to select the environmental driver that best helps capture the local trend data. Time series for these drivers was gathered at the province level for each country. These models and the role of environmental drivers are explained in detail elsewhere [22].

Viral spillover into humans is modelled with a stochastic Susceptible-Exposed-Infectious-Recovered-Susceptible (SEIRS) structure, where the risk of infection depends solely on the prevalence of infection among animals, a transmission coefficient and a factor controlling the excess risk conferred by human activity (i.e. farming or other activities). This means that human infection in our model comprises transmission from livestock to humans, but not human to human.

For each location we calibrate this model to animal and human data (**Table S1**), using a Bayesian framework. Input values and calibrated parameters can be found in **Table S2**. Final calibrated models can be seen in **Figs S1-S3**

### Vaccine candidate profile

Vaccine trial feasibility has multiple associated factors. Here we focus on the trial aspects related to sample size (noted N) and follow-up time under different baseline epidemic conditions.

First, we define the characteristics of a potential effective vaccine candidate against CCHFV following a decision tree for vaccine efficacy trials as proposed by Bellan et al. [23]. The full outcome of this exercise can be found in **Table 1**. In brief, we propose an active controlled individual randomised trial, targeted to groups at high occupational risk in endemic areas, and a trial endpoint that reflects the laboratory-confirmed clinical form of CCHFV disease.

**Table 1.** Randomised controlled trial characteristics for CCHFV. Proposed vaccine trial profile for a CCHFV vaccine efficacy trial of phase III in an endemic area. The characteristics were selected following the InterVax-Tool as proposed by Bellan et al. [23]

| **Trial Characteristic** | **Choice** | **Rationale** |
| --- | --- | --- |
| **Trial population** | Inhabitants of endemic areas | Trials should be carried out in known endemic areas. Also, our previous work on environmental drivers suggests that some climatic conditions can provide a good indication of the time of the year when more CCHF cases can be expected |
| **Risk target** | Farmers/Animal handlers | Our previous research suggests that targeted vaccine campaigns can be more efficient than the general population. Moreover, CCHFV seems to carry a highly defined profile of at-risk population: animal handlers (farmers etc.), butchers, and less importantly health care workers. |
| **Randomization** | Individual randomization | Given that we are looking at spillover transmission, individual randomization is a preferred option and more efficient statistically. |
| **Intervention** | CCHFV vaccine | A safe and effective vaccine formulation should be offered. We assume here a one dose scheme. |
| **Comparator** | Active control | We are proposing an active control which could maximize the benefit of the trial in the community. Meningitis, Hepatitis B or Hepatitis A could be potential options, or even Typhoid conjugate, as long as this vaccine schedule mimics the CCHFV vaccine schedule and blinding can be ensured (i.e., cold-chain, storage). Otherwise a placebo would be the next best option |
| **Primary endpoint** | CCHF clinical case | CCHF is routinely reported and the clinical case is feasible and desirable |
| **Case definition** | Symptomatic case with febrile illness with headache, myalgia, backache, joint or abdominal pain, vomiting, or haemorrhagic manifestations | CCHF has a wide range of symptomatic disease. The more convenient outcome would be the broadly defined clinical cases, confirmed by laboratory. Since onwards transmission from infected humans is expected to be negligible, preventing asymptomatic infection is not immediately relevant. Severe disease can be a secondary outcome. |
| **Case ascertainment** | PCR test confirmation | A PCR test is available and should be used to detect active infection in suspected cases. |
| **Blinding** | Double blinding | Double blind is preferred, and should take priority when assessing if active control or placebo should be used. |

Within this framework we propose a minimum expected vaccine efficacy (VE) of 0.6 (i.e.,60%), which is defined as proposed elsewhere [24],

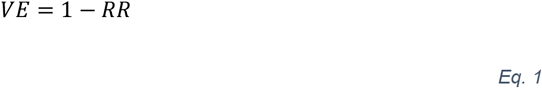

where *RR* is the relative risk of CCHF occurring in the vaccinated arm relative to the control arm. We define *RR* as the ratio of attack rates between study arms, as follows,

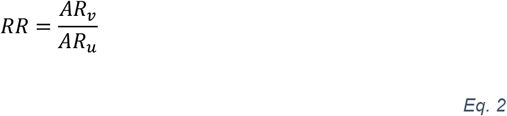

where *AR*_*v*_*sub>*and *AR*_*u*_ are the attack rates of CCHFV over the study period among vaccinees and control groups respectively.

### Sample size calculation

Having defined clinical cases of CCHF as a primary endpoint (Table 1) and an expected vaccine efficacy, we estimate the number of necessary endpoints (*n*_*i*_) to analyse vaccine efficacy under certain conditions of power and significance level (type I error probability). Under this simple approach, we reduce the sample size estimation to a one proportion hypothesis test. We propose a null hypothesis (H_0_) that vaccine efficacy is less than or equal to 30%, and an alternative hypothesis that vaccine efficacy is greater than 60%, the threshold of efficacy previously defined for our vaccine candidate. Next, we solve for *n*_*i*_ from the Chi-Square one proportion test by

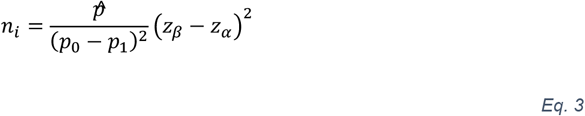

where *p*_0_ and *p*_1_ represent the probability of being a CCHF case when receiving the vaccine under the null and alternative hypothesis, respectively, and *z*_*β*_ and *z*_1 − *α*_ are the Z-score values for the selected power *β* and significance level *α*, respectively.

Here 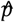 is the effect size (i.e. the magnitude of the difference between *p*_0_ and *p*_1_). We calculate it using the formula for effect size for proportions (Cohen’s *h*) [25]

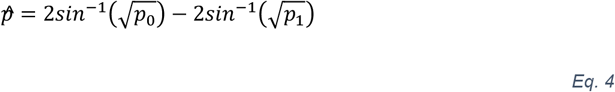

Therefore, for *H*_*1*_ we write

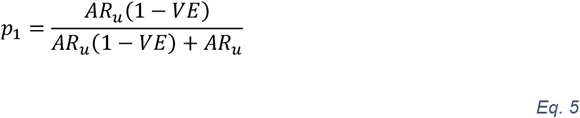

where VE is 0.6 and the attack rate in the control group *AR*_*u*_ is estimated.

### Simulating attack rates

The approach mentioned above requires an estimate of the expected attack rate of symptomatic CCHF among the unvaccinated control group (*AR*_*u*_). We estimate *AR*_*u*_ by reproducing a closed cohort design nested in the previously calibrated transmission model for each country. As mentioned before, the human model follows a SEIRS design, in which the risk of infection is a function of transmission rate *β*_*f*_ (which comprises animal-livestock contact rates and per-contact transmission probabilities), occupational excess risk (e.g., farmers incur a higher risk of infection) and the seroprevalence of CCHFV among livestock. By running instances of the calibrated model (i.e., samples from the posterior density of the calibrated parameters) over this SEIRS cohort we are able to seed the desired susceptible sample size at *t*_*0*_ and follow them up for a period of up to 5 years. The attack rate (*AR*_*u*_) is then easily calculated by

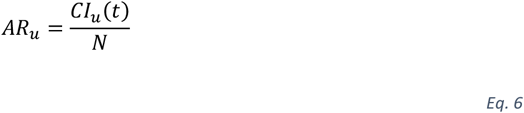

with *CI*_*u*_(*t*) as the cumulative incidence of clinical CCHFV up to time *t*, and *N as* the sample size of susceptible controls at *t*_*0*_. We focus primarily on attack rates of the most common clinical form of the disease but we also take a wider spectrum from infection to fatality in order to assess the feasibility of alternative study endpoints in different settings. Importantly, we assume that, in the context of a randomised controlled trial, CCHF cases are actively ascertained among the suspected cases, therefore reporting capacities in each setting do not play a role in sample size estimation in this case.

The calibrated simulations run over different time periods but all spanning from April 2007 until December 2021. We seed a study cohort in 2008, selecting the month preceding the highest expected seasonal peak of CCHFV cases in each setting: March in Turkey, May in Afghanistan, and September in South Africa. By doing this, we assume first that operationally this minimises the lead-time before accruing the necessary endpoints, and second, that this is a single-dose vaccine, with a month being sufficient time to achieve protection. **Figure 2** shows the simulated incidence, and the different seasonal patterns in the three countries.

**Figure 2.**
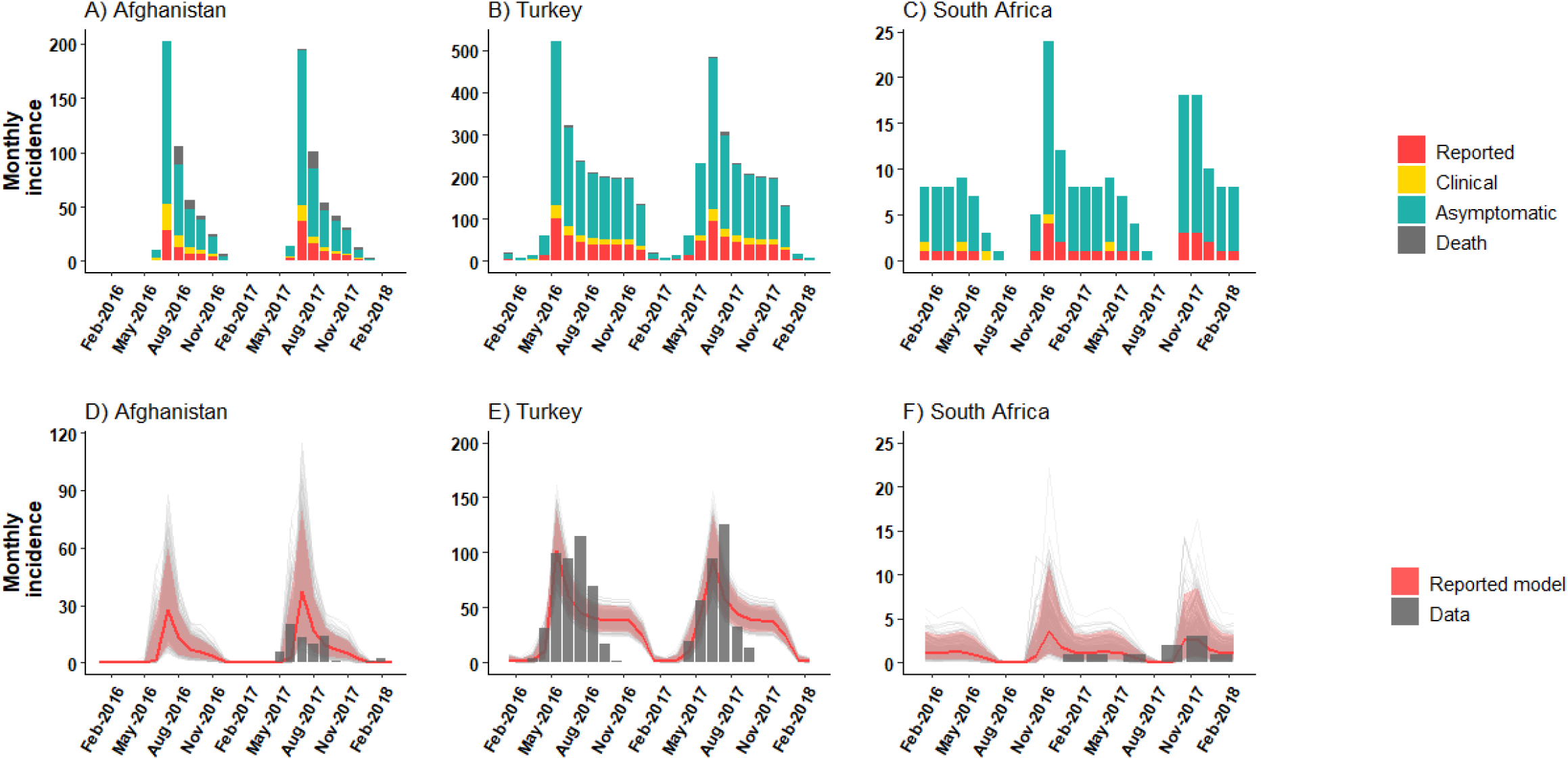
Human CCHF Incidence trajectories in endemic areas. Top panels A-C show the simulated monthly incidence of CCHF cases reported (red), cases not-reported (yellow), asymptomatic infections (green), and fatalities (grey). The bars reflect the median estimate. Bottom row (D-F) show the simulated incidence of reported cases against country-specific data (grey bars). Uncertainty around the median estimate (solid red) is shown in shaded red (95% CrI). Stochastic trajectories of 500 samples from the posterior distribution are shown in light grey.

### Analysis of vaccine trial feasibility

We focus here on two central aspects of trial feasibility, namely, estimated sample size needed to reach the necessary number of study endpoints (*n*_*i*_ in equation 3), and the estimated time to reach those outcomes. We explore the change in these estimates across the locations we model while exploring varying assumptions about the expected profile of VE.

## Results

Transmission models for CCHFV amongst livestock and humans were calibrated to data from endemic areas in Afghanistan, South Africa and Turkey (**Figs S1-S3**). As reported before, saturation deficit is the environmental driver that best describes seasonality and tick activity in Afghanistan [10]. In Turkey and South Africa, soil temperature is a better surrogate marker, according to DIC (See **Fig S4**). However, the relative difference in DIC for saturation deficit and soil temperature is not considered significant. The model calibrations also highlight the different endemic levels of CCHFV spillover transmission into humans in the three countries (**Fig 2**) and seasonal patterns determined by the annual cycles described by the environmental drivers (e.g., soil temperature, saturation deficit) in each location. From these patterns, our selected optimal times for starting trial recruitment in each setting appear to be May, April and October in Afghanistan, Turkey and South Africa, respectively. We estimate that the highest risk of human spillover transmission can be found in Herat, Afghanistan, where the attack rate of CCHF after 6 months of follow-up is 5.5 (CrI 95%, 1.6 – 11. 2) per 1,000 individuals at high risk (Table 2), followed by Turkey with5.1 (CrI 95%, 2.5 - 16.4), and 0.01 (CrI 95%,0.001 - 0.02) in South Africa.

**Table 2.** Estimates of trial design parameters for a CCHVF vaccine in endemic settings in three countries

| **Estimate** | **Afghanistan**<br>Mean (95% Credible interval) | **South Africa**<br>Mean (95% Credible interval) | **Turkey**<br>Mean (95% Credible interval) |
| --- | --- | --- | --- |
| Attack rate of CCHF at 6 months follow-up (per 1,000 high risk population) | 5.5 (1.6 - 11.2) | 0.01 (0.001 - 0.02) | 5.1 (2.5 - 16.4) |
| Attack rate of CCHFV infection at 6 months follow-up (per 1,000 high risk population) | 27.6 (8.2 – 56) | 0.1 (0.01 - 0.2) | 17.1 (8.5 -54.6) |
| CCHFV endpoints to assess vaccine efficacy ($n_i$)<sup>§</sup> | 150 | 150 | 150 |
| Minimum sample size ($N$) to reach clinical endpoints at 6 months | 34,000 (16,750 – 88,725) | NA | 37,000 (13,000 – 77,250) |
| Minimum sample size ($N$) to reach infection endpoints at 6 months | 7,000 (3,275 – 29,725) | NA | 11,000 (3,000 – 23,000) |
| Follow-up time to reach $n_i$ clinical cases<sup>¥</sup> (months) | 5 (2 – 26) | >160 | 6 (2 – 16) |
| Follow-up time to reach $n_i$ clinical cases<sup>¥</sup> (months) | 2 (2 – 3) | 159 (61 – 160) | 2 (2 – 4) |

<sup>§</sup> Assuming a vaccine efficacy target of 60% powered at 90% and 5% significance
<sup>¥</sup> For an assumed sample size of 30,000, 60% powered at 90% and 5% significance

In these settings we assess the number of events necessary to evaluate vaccine efficacy through the vaccine trial proposed in this study. We estimate that for a randomised controlled trial with an expected vaccine efficacy of 60%, with significance level of 5% and powered at 90%, it would be necessary to accrue at least 150 CCHF events. This estimate is the same across all settings. When we increase the expected vaccine efficacy (*H*_*1*_), the effect size of a potential effective vaccine becomes larger, thus fewer endpoints are required to confidently reject the null hypothesis (**Fig 4**). At the same time, a higher target VE results in shorter lead-times to reach these outcomes in all the settings, as expected.

Conducting a trial with the preferred outcome of clinical CCHF cases would be feasible in a setting with transmission levels like those in Herat, Afghanistan, or in the northeast endemic provinces of Turkey, where a sample size of 34,000-37,000 can yield the necessary endpoints in a mean time of ∼ 6 months (**Fig 3, Table 2**). For the modelled endemic areas in South Africa this would not be possible inside the 5 year time window we simulated, even for large sample sizes and target vaccine efficacy over 90%, as seen in **Table 2** and **Fig 4A – C**.

**Figure 3.**
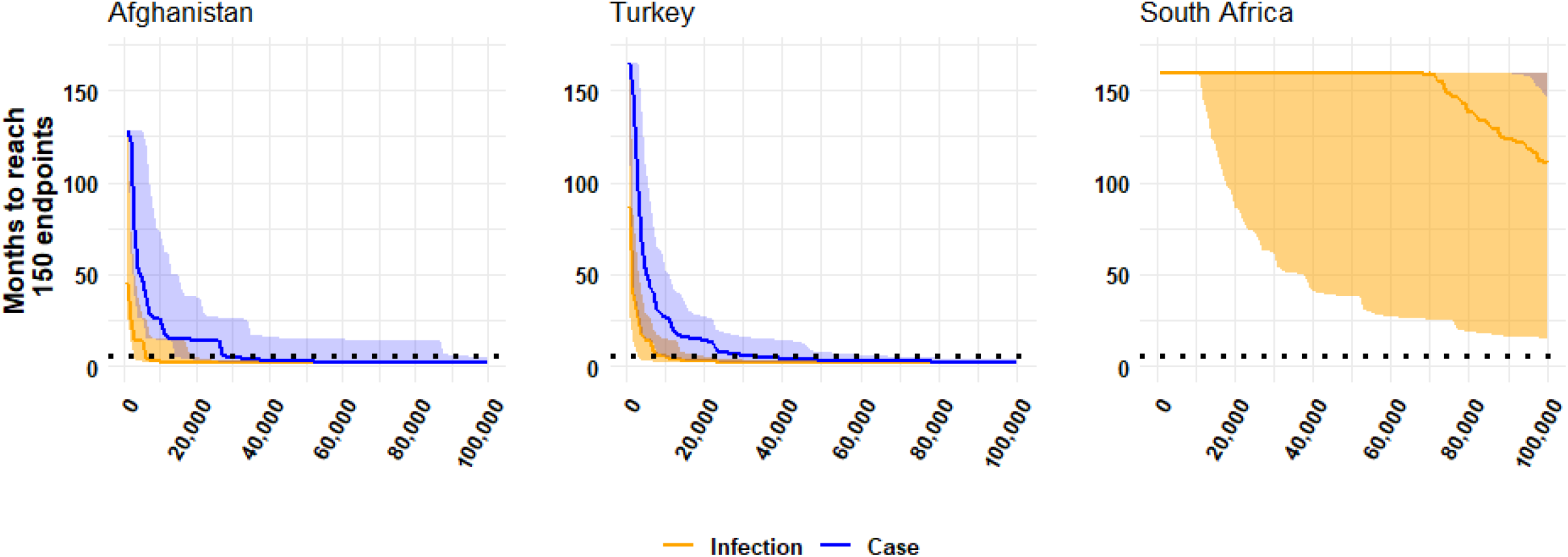
Follow-up-time to reach CCHF endpoint cases. Estimated number of months of trial follow-up necessary to reach trial endpoints (infection in yellow, cases in blue) for a randomised controlled trial with a target vaccine efficacy of 60%. Shaded areas show the uncertainty (95% CrI) in the epidemic model in the simulated cohort. Solid line shows 50% percentile of the sample posterior for these outcomes. A plausible threshold of follow-up time equal to 6 months is shown by the black dotted line.

**Figure 4.**
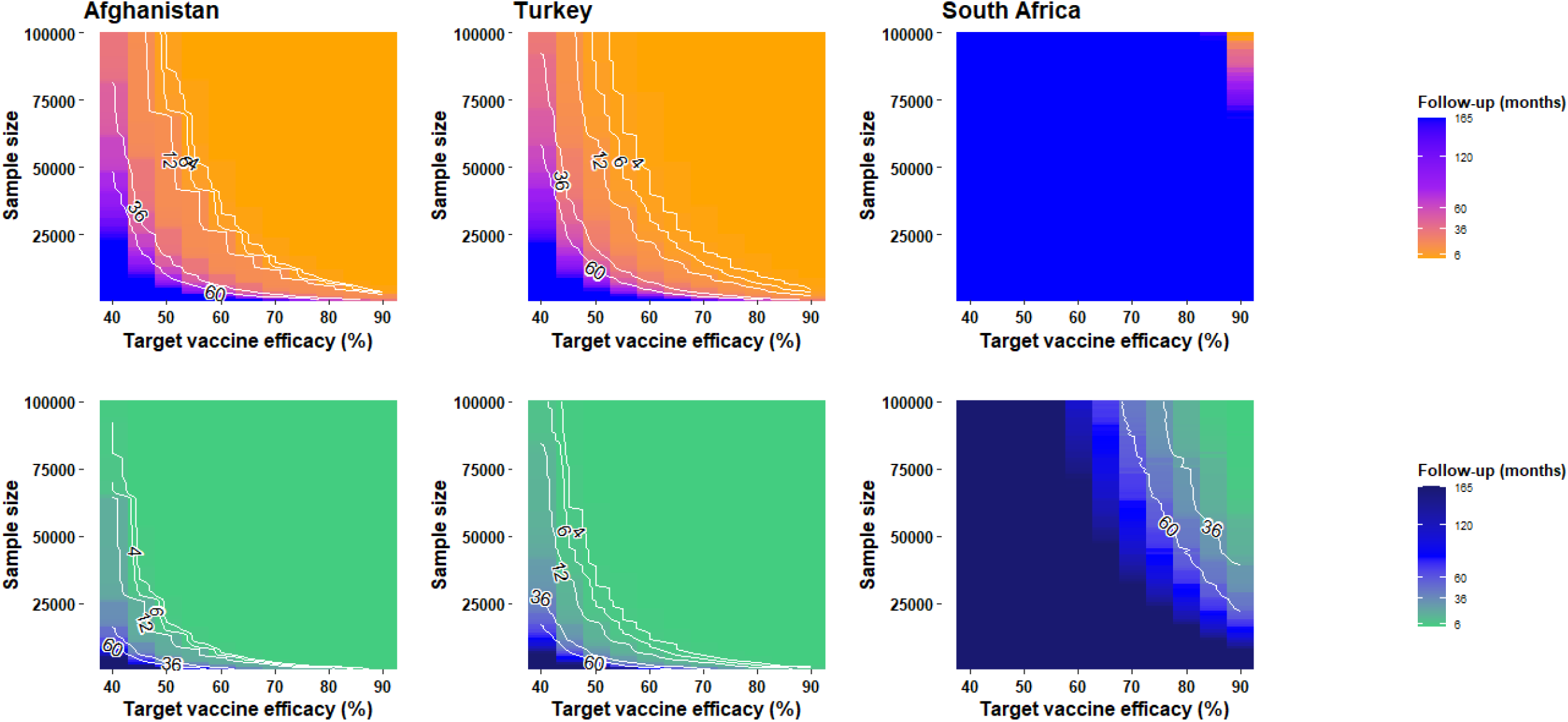
Follow-up time to reach trial endpoints by sample size and target vaccine efficacy in three countries. Heat map and contour lines show months of trial follow-up required under combinations of scenarios of initial sample size and target vaccine efficacy. In top row panels this analysis is performed for a trial scenario where a clinical trial endpoint is selected. In bottom rows, this analysis is for an infection endpoint.

Using CCHFV infection as a primary endpoint (i.e., clinical + paucisymptomatic + asymptomatic), could mean that evaluating vaccine efficacy could be attained in Afghanistan and Turkey at even half the estimated required sample size, in a ∼6 months period (**Table 2**). In South Africa, a trial becomes feasible in this scenario for large sample sizes (>75,000) and target vaccine efficacy of >70% (**Fig 4D-E**).

## Discussion

We have designed and calibrated CCHFV transmission models for livestock and human spillover in endemic areas in Afghanistan, Turkey and South Africa, in order to assess the feasibility of vaccine efficacy trials while accounting for differences in transmission dynamics. The results indicate that a randomised controlled trial to assess VE, targeted to high risk groups (e.g. farmers) with clinical disease as primary outcome, will be feasible if at least 150 events are accrued, under standard conditions of power and confidence. This threshold could be achieved in a mean time of 6 months in Herat, Afghanistan, with a sample size ∼34,000, while in the Kelkit valley, in Turkey, the same could be achieved for a sample size of ∼37,000 and over. Despite a higher volume of CCHF cases reported per year in Turkey, when controlling for population size at risk our attack rates estimates indicate that Afghanistan has a slightly higher risk per individual than Turkey (**Table 2**). By this criterion (i.e., attack rate) Herat would offer a more efficient setting for conducting a trial, yet other operational challenges need to be taken into account when selecting the most convenient study site.

We find that shorter follow-up periods could be possible in these settings if the assumption of a hypothetical 60% vaccine efficacy is increased further. In South Africa, our results show that the very low number of cases might not be enough to conduct this type of trial in a reasonable time-span. We conclude that the requisites of time and sample size for a vaccine trial against CCHFV do vary according to local epidemic conditions. For Turkey and Afghanistan, our results show that adopting a trial endpoint like CCHFV infection (instead of clinical CCHF) will greatly improve the possibility of accruing trial endpoints in less than 6 months and with smaller sample sizes. However, the relevance of powering a CCHFV vaccine trial with such an endpoint is unclear, given that it is expected that the vast majority of cases will arise as either tick-borne or zoonotic transmission. Therefore, the secondary gains of transmission blocking can be considered not relevant for the purpose of designing CCHFV vaccine trials.

In all locations we found that the calibrated model is consistent with a seasonal pattern in CCHFV in humans, strongly driven by environmental factors (i.e., saturation deficit, soil temperature), mirroring the endemic levels simulated among livestock. Our modelling suggests that endemic areas in Turkey appear to have a longer period of active transmission into humans through the year (i.e., April to February, peaking in June), which accounts for the much larger volume of infections observed in our simulations. Crucially, this suggests that Turkey’s high number of cases is not an effect of an established active surveillance system but of higher underlying levels of transmission. In South Africa we reach a similar conclusion from opposing findings: low endemicity is not an effect of case underreporting but of levels of spillover and livestock prevalence. Although it has been suggested that CCHFV has extensively been underestimated in Africa, it is important to note that countries like South Africa and Uganda have a much better established system of surveillance than their neighbouring countries [26].

In Afghanistan our model shows a well-defined cycle of high transmission between May and July, which is in line with the wide climatic range in this area, which drives tick activity seasonality (**Fig 2**). Within this cycle, spillover transmission goes extinct for an extended period before re-emergence occurs in a predictable pattern, without being extensively altered by stochastic effects. This is better exemplified when observing the simulated incidence in South Africa, where despite the very low monthly yield of CCHF cases, the seasonal cycle remains predictable over time without falling into extinction (**Fig 2, Fig S3)**. This is an important insight and also relevant to our aim of assessing vaccine trials for CCHFV, since these dynamics are consistent with a zoonotic infection with established endemicity, indicating that alternative designs like ring vaccination trials used during Ebola outbreaks [27,28] might not be necessary for this disease. Ring vaccination was designed to exploit the clustering of cases within contacts of cases (or their contacts) in a scenario where the emergence of new human infections is highly uncertain. Therefore, in the absence of clear chains of human to human transmission and with a predictable re-emerging pattern in human cases of CCHF, the use of traditional designs like the individual randomised controlled trial is feasible and recommended.

In this analysis we have assumed that these transmission patterns remain unchanged beyond the described dynamics. That is, we simulate trial recruitment and follow-up under calibrated conditions of livestock and spillover transmission. It is, however, plausible that changes in the ecology of the multiple hosts and vectors, or in human activity might result in outbreak events like those observed sporadically in several parts of the world [29–31].

The trial requirements and minimum sample sizes estimated in this analysis do not account for potential operational challenges like attrition of subjects or limitations in test performance during case ascertainment. These are important aspects of trial design and execution and will require further exploration.

Our model does not explicitly incorporate a detailed module for vector cycle and transmission. This is due to the paucity of location-specific data on tick activity and abundance. However, our approach using environmental drivers as a proxy indicator of tick activity, allows us to confidently capture the variation in transmission among livestock and therefore in risk among humans.

In conclusion, our work has assessed for the first time the transmission dynamics of CCHFV in multiple epidemiological settings and the feasibility of conducting phase III vaccine efficacy trials in such locations. This work highlights the epidemiological implications of varying levels of human spillover transmission when defining the necessary minimums for statistical assessment of efficacy in terms of sample size and follow-up time. This work breaks ground for future assessment and establishment of a CCHFV vaccine roadmap.

## Data Availability

All data produced in the present work are contained in the manuscript or available in the references cited in the manuscript

## References

[1] Christova I, Kovacheva O, Georgieva G, Ivanova S, Argirov D. Vaccine against congo-crimean haemorrhagic fever virus-bulgarian input in fighting the disease. Probl Infect Parasit Dis 2010:7–8.

[2] Canakoglu N, Berber E, Tonbak S, Ertek M, Sozdutmaz I, Aktas M, et al. Immunization of knock-out α/β interferon receptor mice against high lethal dose of Crimean-Congo hemorrhagic fever virus with a cell culture based vaccine. PLoS Negl Trop Dis 2015;9. https://doi.org/10.1371/JOURNAL.PNTD.0003579.

[3] Spik K, Shurtleff A, McElroy AK, Guttieri MC, Hooper JW, Schmaljohn C. Immunogenicity of combination DNA vaccines for Rift Valley fever virus, tick-borne encephalitis virus, Hantaan virus, and Crimean Congo hemorrhagic fever virus. Vaccine 2006;24:4657–66. https://doi.org/10.1016/J.VACCINE.2005.08.034.

[4] Farzani TA, Földes K, Ergünay K, Gurdal H, Bastug A, Ozkul A. Immunological Analysis of a CCHFV mRNA Vaccine Candidate in Mouse Models. Vaccines (Basel) 2019;7. https://doi.org/10.3390/VACCINES7030115.

[5] Ghiasi SM, Salmanian AH, Chinikar S, Zakeri S. Mice orally immunized with a transgenic plant expressing the glycoprotein of Crimean-Congo hemorrhagic fever virus. Clin Vaccine Immunol 2011;18:2031–7. https://doi.org/10.1128/CVI.05352-11.

[6] Estrada-Peña A, Vatansever Z, Gargili A, Ergönul Ö. The trend towards habitat fragmentation is the key factor driving the spread of Crimean-Congo haemorrhagic fever. Epidemiol Infect 2010;138:1194–203. https://doi.org/10.1017/S0950268809991026.

[7] Estrada-Peña A, Vatansever Z, Gargili A, Buzgan T. An early warning system for Crimean-Congo haemorrhagic fever seasonality in Turkey based on remote sensing technology. Geospat Health 2007;2:127–35. https://doi.org/10.4081/GH.2007.261.

[8] Vescio FM, Busani L, Mughini-Gras L, Khoury C, Avellis L, Taseva E, et al. Environmental correlates of Crimean-Congo haemorrhagic fever incidence in Bulgaria. BMC Public Health 2012;12. https://doi.org/10.1186/1471-2458-12-1116.

[9] World Health Organization. An R&D blueprint for action to prevent epidemics. Geneva 2016. https://www.who.int/blueprint/about/r_d_blueprint_plan_of_action.pdf (accessed November 23, 2021).

[10] Vesga JF, Clark MHA, Ayazi E, Apolloni A, Leslie T, Edmunds WJ, et al. Transmission dynamics and vaccination strategies for Crimean-Congo haemorrhagic fever virus in Afghanistan: A modelling study. PLoS Negl Trop Dis 2022;16:e0010454. https://doi.org/10.1371/JOURNAL.PNTD.0010454.

[11] Sahak M, Arifi F, Saeedzai S. Descriptive epidemiology of Crimean-Congo Hemorrhagic Fever (CCHF) in Afghanistan: Reported cases to National Surveillance System, 2016-2018. Int J Infect Dis 2019;88:135–40. https://doi.org/10.1016/J.IJID.2019.08.016.

[12] Mofleh J, Ahmad Z. Crimean-Congo haemorrhagic fever outbreak investigation in the Western Region of Afghanistan in 2008. Eastern Mediterranean Health Journal = La Revue de Sante de La Mediterranee Orientale = Al-Majallah al-Sihhiyah Li-Sharq al-Mutawassit 2012;18:522–6. https://doi.org/10.26719/2012.18.5.522.

[13] Niazi A, Jawad M, Amirnajad A, Durr P, Williams D. Crimean-Congo Hemorrhagic Fever, Herat Province, Afghanistan, 2017. Emerg Infect Dis 2019;25:1596–8. https://doi.org/10.3201/EID2508.181491.

[14] Mustafa ML, Ayazi E, Mohareb E, Yingst S, Zayed A, Rossi CA, et al. Crimean-Congo Hemorrhagic Fever, Afghanistan, 2009. Emerging Infectious Diseases 2011;17:1940. https://doi.org/10.3201/EID1710.110061.

[15] National institute for communicable diseases. Crimean-Congo haemorrhagic fever. Communique (Wash DC) 2022;19:3.

[16] Fisher-Hoch SP, McCormick JB, Swanepoel R, Middelkoop A van, Harvey S, Kustner HG v. Risk of Human Infections with Crimean-Congo Hemorrhagic Fever Virus in a South African Rural Community. The American Journal of Tropical Medicine and Hygiene 1992;47:337–45. https://doi.org/10.4269/AJTMH.1992.47.337.

[17] Msimang V, Weyer J, Roux C le, Kemp A, Burt FJ, Tempia S, et al. Risk factors associated with exposure to Crimean-Congo haemorrhagic fever virus in animal workers and cattle, and molecular detection in ticks, South Africa. PLoS Negl Trop Dis 2021;15. https://doi.org/10.1371/JOURNAL.PNTD.0009384.

[18] Yilmaz GR, Buzgan T, Irmak H, Safran A, Uzun R, Cevik MA, et al. The epidemiology of Crimean-Congo hemorrhagic fever in Turkey, 2002–2007. International Journal of Infectious Diseases 2009;13:380–6. https://doi.org/10.1016/J.IJID.2008.07.021.

[19] Monsalve-Arteaga L, Alonso-Sardón M, Bellido JLM, Santiago MBV, Lista MCV, Abán JL, et al. Seroprevalence of crimean-congo hemorrhagic fever in humans in the world health organization european region: A systematic review. PLoS Neglected Tropical Diseases 2020;14:1–15. https://doi.org/10.1371/journal.pntd.0008094.

[20] Leblebicioglu H, Ozaras R, Irmak H, Sencan I. Crimean-Congo hemorrhagic fever in Turkey: Current status and future challenges. Antiviral Research 2016;126:21–34. https://doi.org/10.1016/J.ANTIVIRAL.2015.12.003.

[21] Turkey records 13 Crimean-Congo Hemorrhagic Fever deaths year to date -Outbreak News Today n.d. http://outbreaknewstoday.com/turkey-records-13-crimean-congo-hemorrhagic-fever-deaths-year-to-date-37034/ (accessed November 4, 2021).

[22] Vesga JF, Clark MHA, Ayazi E, Apolloni A, Leslie T, Edmunds WJ, et al. Transmission dynamics and vaccination strategies for Crimean-Congo haemorrhagic fever virus in Afghanistan: a modelling study. MedRxiv 2022:2022.01.20.22269558. https://doi.org/10.1371/journal.pntd.0010454.

[23] Bellan SE, Eggo RM, Gsell PS, Kucharski AJ, Dean NE, Donohue R, et al. An online decision tree for vaccine efficacy trial design during infectious disease epidemics: The InterVax-Tool. Vaccine 2019;37:4376–81. https://doi.org/10.1016/J.VACCINE.2019.06.019.

[24] O’neill RT. On sample sizes to estimate the protective efficacy of a vaccine. Stat Med 1988;7:1279–88. https://doi.org/10.1002/SIM.4780071208.

[25] Cohen J. Statistical power analysis for the behavioral sciences (2nd ed.). Hillsdale, NJ: Lawrence Earlbaum Associates. Lawrence Earlbaum Associates 1988:286.

[26] Temur AI, Kuhn JH, Pecor DB, Apanaskevich DA, Keshtkar-Jahromi M. Epidemiology of Crimean-Congo Hemorrhagic Fever (CCHF) in Africa-Underestimated for Decades. Am J Trop Med Hyg 2021;104:1978–90. https://doi.org/10.4269/AJTMH.20-1413.

[27] The ring vaccination trial: a novel cluster randomised controlled trial design to evaluate vaccine efficacy and effectiveness during outbreaks, with special reference to Ebola. BMJ 2015;351:h3740. https://doi.org/10.1136/BMJ.H3740.

[28] Henao-Restrepo AM, Longini IM, Egger M, Dean NE, Edmunds WJ, Camacho A, et al. Efficacy and effectiveness of an rVSV-vectored vaccine expressing Ebola surface glycoprotein: interim results from the Guinea ring vaccination cluster-randomised trial. Lancet 2015;386:857–66. https://doi.org/10.1016/S0140-6736(15)61117-5.

[29] Mirembe BB, Musewa A, Kadobera D, Kisaakye E, Birungi D, Eurien D, et al. Sporadic outbreaks of crimean-congo haemorrhagic fever in Uganda, July 2018-January 2019. PLOS Neglected Tropical Diseases 2021;15:e0009213. https://doi.org/10.1371/JOURNAL.PNTD.0009213.

[30] Kizito S, Okello PE, Kwesiga B, Nyakarahuka L, Balinandi S, Mulei S, et al. <em>Notes from the Field</em>: Crimean-Congo Hemorrhagic Fever Outbreak — Central Uganda, August– September 2017. MMWR Morbidity and Mortality Weekly Report 2019;67:646–7. https://doi.org/10.15585/MMWR.MM6722A6.

[31] Karti SS, Odabasi Z, Korten V, Yilmaz M, Sonmez M, Caylan R, et al. Crimean-Congo Hemorrhagic Fever in Turkey - Volume 10, Number 8—August 2004 - Emerging Infectious Diseases journal - CDC. Emerging Infectious Diseases 2004;10:1379–84. https://doi.org/10.3201/EID1008.030928.

