## Supporting information for "Vaccine efficacy trials for Crimean-Congo haemorrhagic fever: insights from modelling different epidemiological settings"

**Table S1: Target data used in model calibration**

| Calibration target | Description | Year(s) | Source |
|---|---|---|---|
| **Afghanistan** | | | |
| Livestock seroprevalence of CCHFV | Age stratified IgG seroprevalence from a serosurvey in Herat (n=132) | 2009 | Mustafa *et al*. 2011[1] |
| Human seroprevalence of CCHFV | IgG seroprevalence in humans by occupation in Herat (n=330) | 2009 | Mustafa *et al*. 2011[1] |
| Monthly Human CCHFV cases reported | Reported human cases in Herat. Cases in 2018 at national level and assumed that ~62% are from Herat according to Niazi et al.[2] | 2008, 2017, 2018 | Mofleh et al[3]<br>Niazi et al[2]<br>Sahak et al[4] |
| Yearly Human CCHFV cases reported | Yearly aggregated cases reported nationally. Assumed that ~62% are from Herat according to Niazi et al.[2] | 2009, 2010, 2010, 2011, 2012, 2013, 2014, 2015, 2016 | Niazi et al[2]<br>Sahak et al[4] |
| Yearly Human CCHFV fatalities reported | Yearly aggregated deaths reported nationally. Assumed that ~62% are from Herat according to Niazi et al.[2] | 2009, 2010, 2010, 2011, 2012, 2013, 2014, 2015, 2016 | Niazi et al[2]<br>Sahak et al[4] |
| **Turkey** | | | |
| Livestock seroprevalence of CCHFV | IgG seroprevalence among livestock | 2013, 2017 | Ozan et al., [5]<br>Tekelioglu et al [6] |
| Human seroprevalence of CCHFV | IgG seroprevalence in humans by occupation | 2006,2009,2010 | Gozel et al [7], Bodur et al[8], Koksal[9] |
| Monthly Human CCHFV cases reported | Reported human cases in five provinces in Northeast Turkey | Jan 2004 to Dec 2017 | Ak et al., [10] |
| **South Africa** | | | |
| Livestock seroprevalence of CCHFV | Age stratified IgG seroprevalence from a serosurvey | 2017 | Msimang et al[11] |
| Human seroprevalence of CCHFV | IgG seroprevalence I humans by occupation i | 2017 | Msimang et al[11] |
| Monthly Human CCHFV cases reported | Reported human cases in three states in South Africa | Jan 2000 to Dec 2017 | NICD South Africa [12] |

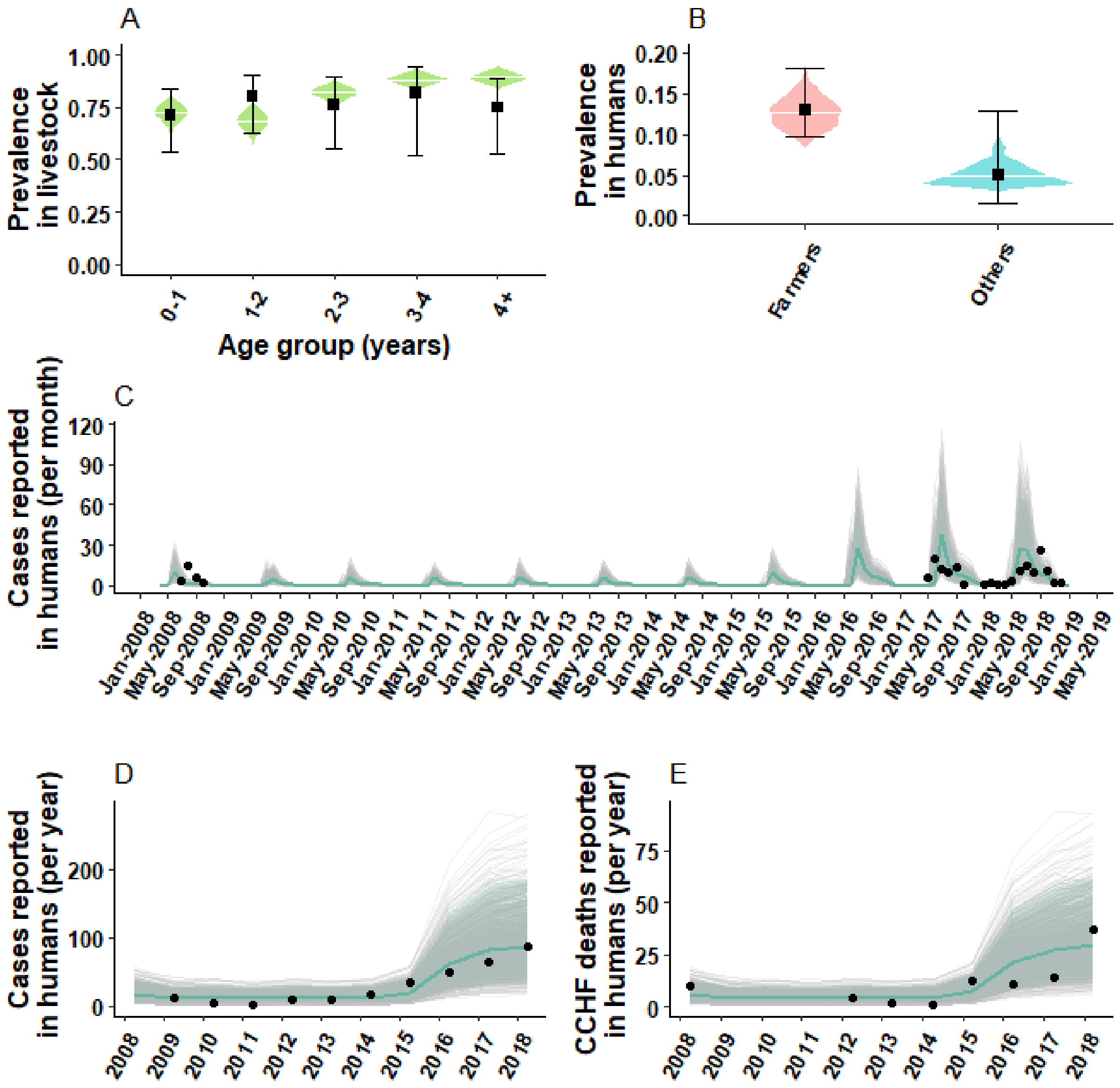


**Figure S1: Model trajectories against calibration target data in Herat, Afghanistan:** Panel A shows the age stratified simulated CCHFV IgG prevalence among livestock (green density plot), with the median estimate (white horizontal line), against IgG prevalence data for the same age groups (black square shows the mean and error bars the 95%CI). Panel B shows the posterior density and median estimate of IgG prevalence for the population of farmers and other occupations (density plots pink and blue) against IgG prevalence data. We take the prevalence estimate to match the dates of data collection as reported. Panel C shows stochastic model trajectories (grey lines) for monthly incident CCHFV human cases reported. In shaded pale grey, the 95% CrI and in solid blue, the median estimate. In black dots, monthly incident cases reported in two separate CCHF outbreaks in Herat: in 2008 as reported by Mofleh et al [3], and 2017 -2018 as reported by Niazi et al, and Sahak et al [2,4]. In Panels D and E, yearly CCHF cases and deaths reported from Herat, against data (black) as reported by Sahak et al.

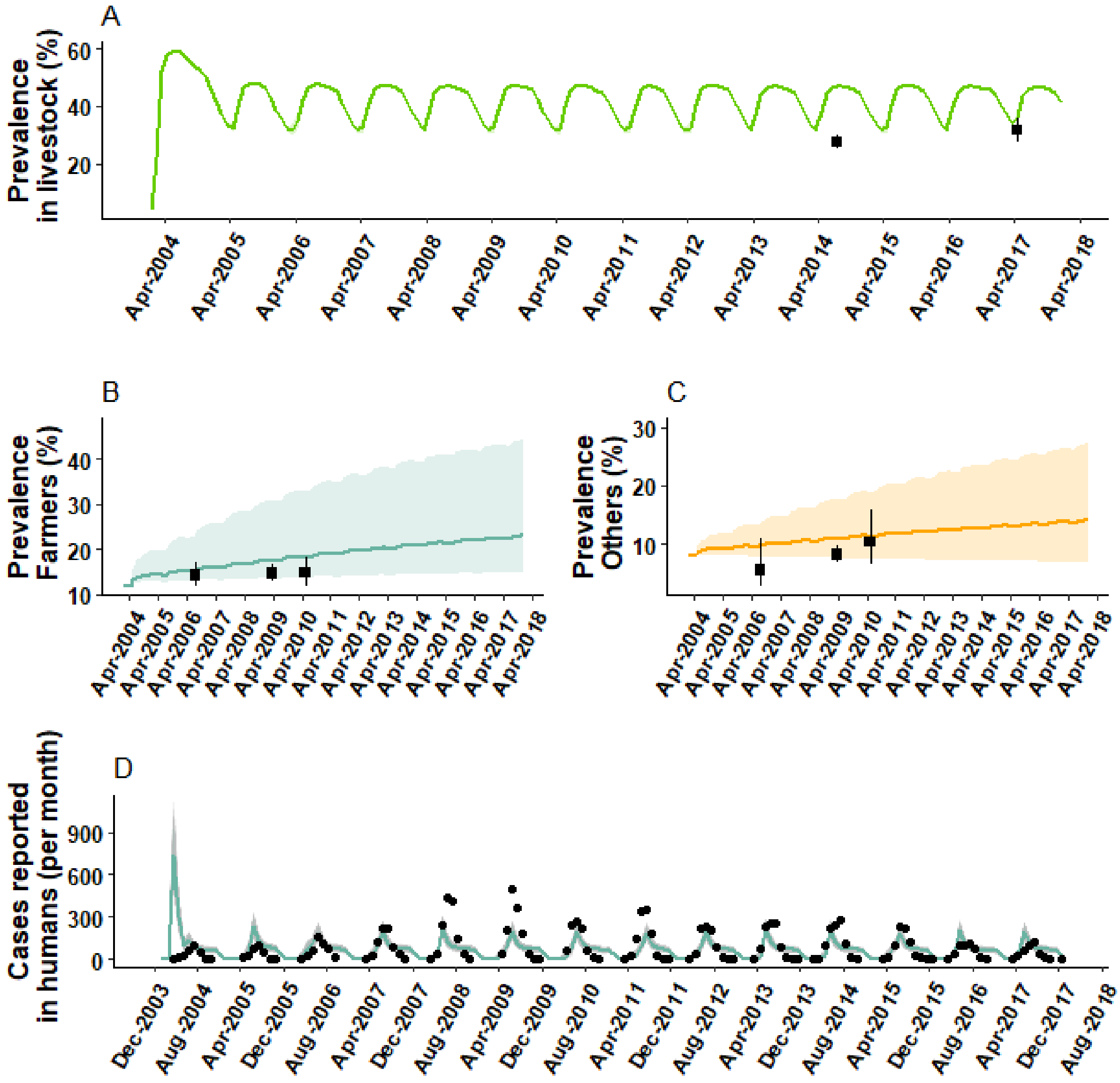


**Figure S2: Model trajectories against calibration target data in northeast Turkey:** Panel A shows the simulated CCHFV IgG prevalence among livestock with the median estimate (green line), against IgG prevalence data (black square shows the mean and error bars the 95%CI). Panel B and C shows the posterior density and median estimate of IgG prevalence over time for the population of farmers and other occupations against IgG prevalence data. We take the prevalence estimate to match the dates of data collection as reported. Panel D shows stochastic model trajectories (grey lines) for monthly incident CCHFV human cases reported. In shaded pale grey, the 95% CrI and in solid blue, the median estimate. In black dots, monthly incident cases reported.

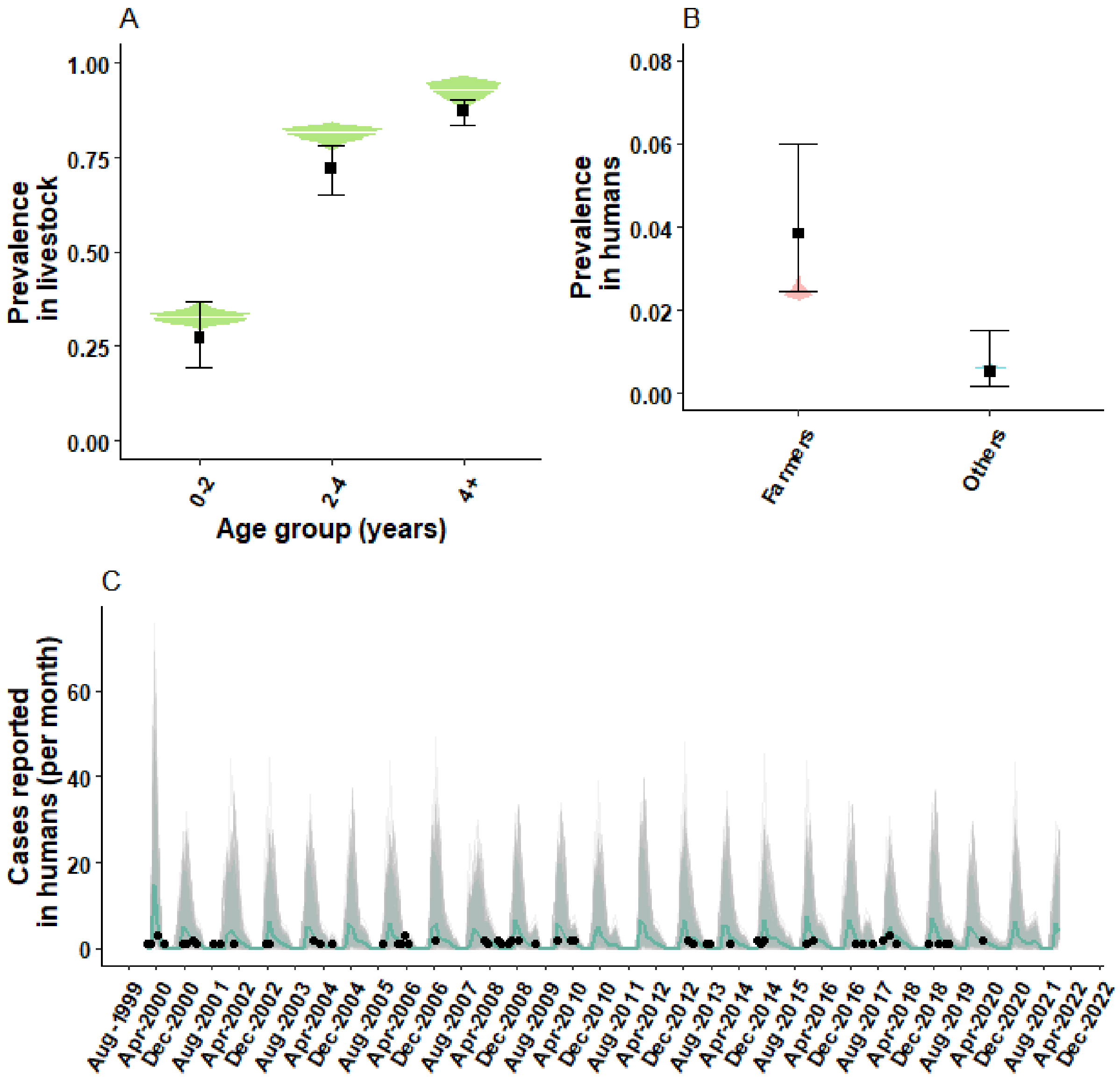


**Figure S3: Model trajectories against calibration target data in three provinces in South Africa:** Panel A shows the age stratified simulated CCHFV IgG prevalence among livestock (green density plot), with the median estimate (white horizontal line), against IgG prevalence data for the same age groups (black square shows the mean and error bars the 95%CI). Panel B shows the posterior density and median estimate of IgG prevalence for the population of farmers and other occupations against IgG prevalence data. We take the prevalence estimate to match the dates of data collection as reported. Panel C shows stochastic model trajectories (grey lines) for monthly incident CCHFV human cases as reported by NICD[12]. In shaded pale grey, the 95% CrI and in solid blue, the median estimate. In black dots, monthly incident cases reported.

**Table S2: Model parameters**

| Parameter description | Notation | Input Values/Estimated* | | | Sources |
|---|---|---|---|---|---|
| | | Afghanistan | Turkey | South Africa | |
| ***Livestock*** | | | | | |
| Duration of infectiousness in livestock | $D_{iL}$ | 7 days | | | Gonzalez et al., 1998[13] |
| Duration of colostrum acquired immunity (months) | $D_{aL}$ | 8.3 (CrI 95% 2-10) | 11.5 (CrI 95% 11-13) | 3.5 (CrI 95% 1.1-11.1) | Estimated |
| Mean time to loss of immunity in adult livestock (months) | $D_{mL}$ | 52 (CrI 95% 46-76) | 38 (CrI 95% 35-44) | 71 (CrI 95% 37-100) | Estimated |
| Proportion of livestock immune at time 0 by age¥ group *a* | $R_a(t)$ | $R_a(t) = \begin{cases} 0.29 & for a = 1 \\ 0.48 & for a = 2 \\ 0.8 & for a = 3 \\ 0.87 & for a = 4 \\ 0.87 & for a = 5 \end{cases}$ | | | Barthel et al., 2014[14] |
| ***Humans*** | | | | | |
| Duration of latent period in humans | $D_{lH}$ | 4 days | | | Bente et al., 2013[15] |
| Duration of infectiousness in humans | $D_{iH}$ | 9 days | | | Fillâtre et al., 2019[16] |

| | | | | | |
|---|---|---|---|---|---|
| Duration of immunity in humans | $D_{mH}$ | 3650 days | | | Assumption |
| Fraction of human infection resulting in a clinical case | $\phi$ | 0.31 (CrI 95% 0.28-0.33) | 0.16 (CrI 95% 0.10-0.30) | 0.19 (CrI 95% 0.10-0.38) | Estimated |
| Proportion of farmers immune at time 0 | $p_F$ | 0.1333 | 0.1333 | 0.05 (assumption) | Mustafa et al., 2011[1] |
| Proportion of others immune at time 0 | $p_O$ | 0.0469 | 0.0469 | 0.02 (assumption) | Mustafa et al., 2011[1] |
| Case fatality rate of CCHF | $CFR_{cchfv}$ | 33% | 25% | 5% | Niazi et al., 2019[2]; NICD [12]; Yilmaz et al [17] |
| ***Demographics*** | | | | | |
| Livestock population size | $N_L$ | 15,193 | 356,981 | 2,800,909 | FAO 2008 [18]; Turkstat[19]; DLRRD[20] |
| Livestock ageing factor (1/months) | $\delta$ | 1/12 | | | Assumption |

| | | | | | |
|---|---|---|---|---|---|
| Livestock monthly death rate | μ | $\mu_a = \begin{cases} 0.0761 & for a = 1 \\ 0.0743 & for a = 2 \\ 0.0746 & for a = 3 \\ 0.0744 & for a = 4 \\ 0.0747 & for a = 5 \end{cases}$ | | | See supplementary material in Vesga et al [21] |
| Population size - Farmers | $N_F$ | 7,614 | 173,622 | 637,383 | USAID 2008 |
| Population size - Other occupations | $N_O$ | 17,768 | 422,763 | 742,234 | USAID 2008 |
| Life expectancy - humans | $L_H$ | 61.5 years | 64 years | 77 years | World bank 2008-2014[22] |
| Monthly birth rate humans | $b_H$ | 1/ (12* $L_H$) | | | Assumption |
| Monthly birth rate in livestock | $b_L$ | μ | | | Assumption |
| ***Viral transmission parameters*** | | | | | |
| Between livestock transmission temperature dependent | A | 0.33 (CrI 95% 0.2-0.4) | 3 (CrI 95% 4-5) | 0.46 (CrI 95% 0.37-0.6) | Estimated |
| Transmission rate from livestock to farmers | $\beta_F$ | 0.28 (CrI 95% 0.15-0.34) | 0.12 (CrI 95% 0.01-0.38) | 0.75 (CrI 95% 0.12-5.1) | Estimated |
| Other occupations relative transmission factor(relative to | O | 0.3 (CrI 95% 0.1-0.5) | 0.43 (CrI 95% 0.04-0.95) | 0.34 (CrI 95% 0.01-0.95) | Estimated |

| | | | | | |
|---|---|---|---|---|---|
| farmers) | | | | | |
| Transmission rate from livestock to other occupations | $\beta_o$ | $O\beta_F$ | | | Assumption |

**Table notes**

**Estimated values represent the posterior mean and 95% CrI for the best most parsimonious model, i.e., saturation deficit obtained during calibration (see section S3 Text* [21] *for calibration details) .*

¥ Livestock age stratification groups where *a*=1 reflects 0 to 12 months; *a*=2 for 13 to 24 months; *a*=3 for 25 to 36 months; *a*=4 for 37 to 48 months, *a*=5 for 48 months and older

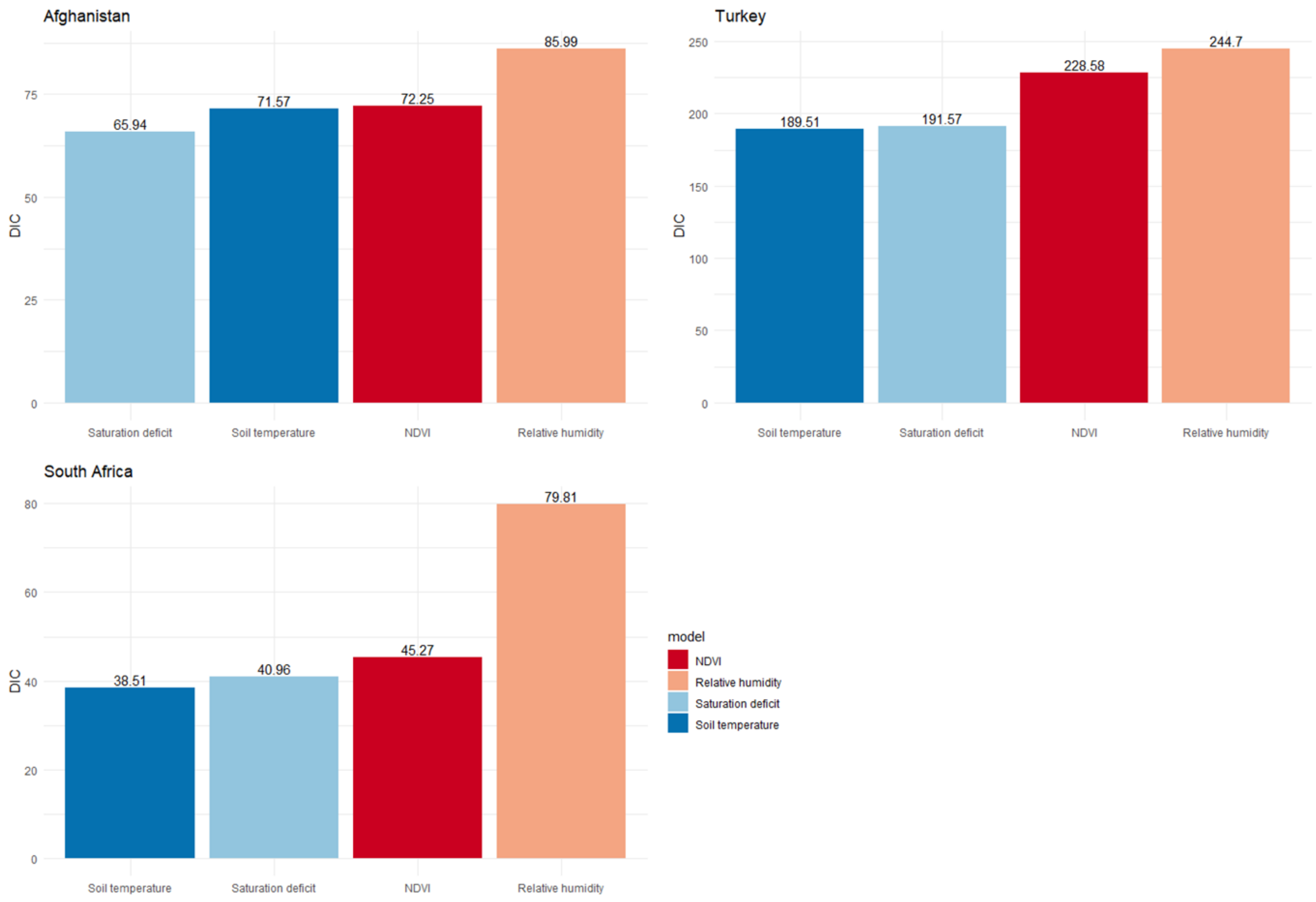


**Figure S4: Deviance information criterion (DIC) for CCHFV transmission models** with different environmental drivers as proxy markers of tick activity. In South Africa and Turkey, soil temperature shows the smallest DIC, while in Afghanistan it is saturation deficit the best performing model.

### References

[1] Mustafa ML, Ayazi E, Mohareb E, Yingst S, Zayed A, Rossi CA, et al. Crimean-Congo Hemorrhagic Fever, Afghanistan, 2009. Emerging Infectious Diseases 2011;17:1940. https://doi.org/10.3201/EID1710.110061.

[2] Niazi A, Jawad M, Amirnajad A, Durr P, Williams D. Crimean-Congo Hemorrhagic Fever, Herat Province, Afghanistan, 2017. Emerg Infect Dis 2019;25:1596–8. https://doi.org/10.3201/EID2508.181491.

[3] Mofleh J, Ahmad Z. Crimean-Congo haemorrhagic fever outbreak investigation in the Western Region of Afghanistan in 2008. Eastern Mediterranean Health Journal = La Revue de Sante de La Mediterranee Orientale = Al-Majallah al-Sihhiyah Li-Sharq al-Mutawassit 2012;18:522–6. https://doi.org/10.26719/2012.18.5.522.

[4] Sahak M, Arifi F, Saeedzai S. Descriptive epidemiology of Crimean-Congo Hemorrhagic Fever (CCHF) in Afghanistan: Reported cases to National Surveillance System, 2016-2018. Int J Infect Dis 2019;88:135–40. https://doi.org/10.1016/J.IJID.2019.08.016.

[5] Ozan E, Ozkul A. Investigation of Crimean-Congo hemorrhagic fever virus in ruminant species slaughtered in several endemic provinces in Turkey. Arch Virol 2020;165:1759–67. https://doi.org/10.1007/S00705-020-04665-9.
[6] Seroepidemiological survey of the Crimean-Congo Hemorrhagic Fever Virus (CCHFV) infection amongst domestic ruminants in Adana province, East Mediterranean, Turkey n.d. https://doi.org/10.31797/vetbio.997150.
[7] Gozel MG, Dokmetas I, Oztop AY, Engin A, Elaldi N, Bakir M. Recommended precaution procedures protect healthcare workers from Crimean-Congo hemorrhagic fever virus. Int J Infect Dis 2013;17:e1046–50. https://doi.org/10.1016/J.IJID.2013.05.005.
[8] Bodur H, Akinci E, Ascioglu S, Öngürü P, Uyar Y. Subclinical Infections with Crimean-Congo Hemorrhagic Fever Virus, Turkey. Emerging Infectious Diseases 2012;18:640. https://doi.org/10.3201/EID1804.111374.
[9] Koksal I, Yilmaz G, Aksoy F, Erensoy S, Aydin H. The seroprevalance of Crimean-Congo haemorrhagic fever in people living in the same environment with Crimean-Congo haemorrhagic fever patients in an endemic region in Turkey. Epidemiol Infect 2014;142:239–45. https://doi.org/10.1017/S0950268813001155.
[10] Ak, Ergönül, Gönen M. A prospective prediction tool for understanding Crimean–Congo haemorrhagic fever dynamics in Turkey. Clinical Microbiology and Infection 2020;26:123.e1-123.e7. https://doi.org/10.1016/J.CMI.2019.05.006.
[11] Msimang V, Weyer J, Roux C le, Kemp A, Burt FJ, Tempia S, et al. Risk factors associated with exposure to Crimean-Congo haemorrhagic fever virus in animal workers and cattle, and molecular detection in ticks, South Africa. PLoS Negl Trop Dis 2021;15. https://doi.org/10.1371/JOURNAL.PNTD.0009384.
[12] National Institute for Communicable Diseases. Crimean-Congo haemorrhagic fever. 2020.
[13] Gonzalez J, Camicas J, Cornet J, virology MW-R in, 1998 undefined. Biological and clinical responses of West African sheep to Crimean-Congo haemorrhagic fever virus experimental infection. Elsevier n.d. https://doi.org/10.1016/S0923-2516(99)80013-2.
[14] Barthel R, Mohareb E, Younan R, Gladnishka T, Kalvatchev N, Moemen A, et al. Seroprevalance of Crimean–Congo haemorrhagic fever in Bulgarian livestock. Taylor & Francis 2014;28:540–2. https://doi.org/10.1080/13102818.2014.931685.
[15] Bente DA, Forrester NL, Watts DM, McAuley AJ, Whitehouse CA, Bray M. Crimean-Congo hemorrhagic fever: history, epidemiology, pathogenesis, clinical syndrome and genetic diversity. Antiviral Research 2013;100:159–89. https://doi.org/10.1016/J.ANTIVIRAL.2013.07.006.
[16] Fillâtre P, Revest M, Tattevin P. Crimean-Congo hemorrhagic fever: An update. Medecine et Maladies Infectieuses 2019;49:574–85. https://doi.org/10.1016/J.MEDMAL.2019.09.005.
[17] Yilmaz GR, Buzgan T, Irmak H, Safran A, Uzun R, Cevik MA, et al. The epidemiology of Crimean-Congo hemorrhagic fever in Turkey, 2002–2007. International Journal of Infectious Diseases 2009;13:380–6. https://doi.org/10.1016/J.IJID.2008.07.021.
[18] FAO. Afghanistan national livestock census 2002-2003. Rome: 2008.
[19] Turkish Statistical Institute (TURKSTAT) n.d. https://www.tuik.gov.tr/Home/Index (accessed May 25, 2022).
[20] Department of Agriculture, Land Reform and Rural Development > Home > Crop Estimates > Statistical Information > Livestock n.d. https://www.dalrrd.gov.za/Home/Crop-Estimates/Statistical-Information/Livestock (accessed May 25, 2022).

[21] Vesga JF, Clark MHA, Ayazi E, Apolloni A, Leslie T, Edmunds WJ, et al. Transmission dynamics and vaccination strategies for Crimean-Congo haemorrhagic fever virus in Afghanistan: A modelling study. PLoS Negl Trop Dis 2022;16:e0010454. https://doi.org/10.1371/JOURNAL.PNTD.0010454.
[22] World Bank. Fertility rate, total (births per woman) - Mexico | Data 2019.
